# Survival-Inferred Fragility of Statistical Significance in Phase III Oncology Trials

**DOI:** 10.1101/2025.01.11.25320398

**Authors:** Alexander D. Sherry, Yufei Liu, Pavlos Msaouel, Timothy A. Lin, Alex Koong, Christine Lin, Joseph Abi Jaoude, Roshal R. Patel, Ramez Kouzy, Molly B. El-Alam, Avital M. Miller, Mohannad Owiwi, Jonathan Ofer, David Bomze, Zachary R. McCaw, Tomer Meirson, Ethan B. Ludmir

## Abstract

**Background:** Statistical significance currently defines superiority in phase III oncology trials. However, this practice is increasingly questioned. Here, we estimated the fragility of phase III oncology trials.

**Methods:** Using Kaplan-Meier curves for the primary endpoints of 230 two-arm superiority phase III oncology trials, we reconstructed data for individual patients. We estimated the survival-inferred fragility index (SIFI) by iteratively flipping the best responder from the experimental arm to the control arm (SIFI_B_) until the interpretation was changed according to the significance threshold of each trial. Severe fragility was defined by SIFI *≤*1%.

**Results:** This study included 230 trials enrolling 184,752 patients. The median number of patients required to change trial interpretation was 8 (interquartile range, 4 to 19) or 1.4% (interquartile range, 0.7% to 3%) per SIFI_B_. Estimations of SIFI by multiple methods were largely consistent. For trials with an overall survival primary endpoint, the median SIFI_B_ was 1% (IQR, 0.5% to 1.9%). Severe fragility was found in 87 trials (38%). As a continuous statistic, the original *P* value—but not its binary significance interpretation—was associated with fragility and severe fragility. Trials with subsequent FDA approval had lower odds of severe fragility. Lastly, the underlying survival model had differential effects on SIFI estimation.

**Conclusions:** Even among phase III oncology trials, which directly inform patient care, changes in the outcomes of few patients are often sufficient to change statistical significance and trial interpretation. These findings imply that current definitions of statistical significance used in phase III oncology are inadequate to identify replicable findings.

## INTRODUCTION

Superiority interpretations in phase III oncology trials are currently governed by statistical significance.^1^ Statistical significance is defined by achieving a *P* value in the survival model less than or equal to a predefined threshold.^2,3^ Although statistical significance is essentially universal in phase III oncology research, the robustness of statistical significance as the chief determinant of treatment superiority has been increasingly questioned.^4,5^ Statistical significance assigns diametrically opposing interpretations to *P* of 0.049 and *P* of 0.051 (when significance is defined as *P* < 0.05, the usual convention), but *P* is a continuous statistic, which can vary stochastically from sample to sample. The information encoded by *P* of 0.049 and 0.051 is nearly identical.^6–10^

To quantify the robustness of statistical significance, the survival-inferred fragility index (SIFI) has been proposed.^11,12^ The SIFI is defined as the minimum number of individual patient outcomes needed to change the interpretation of statistical significance.

Because oncology trials investigate survival, which is a composite of both the timing and occurrence of the event (i.e., alive or deceased), reliable estimation of fragility in oncology research also requires a concurrent evaluation of both patient events and the timing of those events.^13,14^ Thus, to understand fragility in oncology, data for individual patients is required, which has been a major limitation in a field where patient-level data are rarely shared.^15^

Previously, Bomze and colleagues reconstruction survival data for individual patients in 45 phase III trials testing immunotherapy strategies and found a median SIFI of only 5 patients.^16^ Building on this important work, we aimed to estimate fragility among a superset of 230 phase III oncology trials. The purpose of the present study was to provide an updated and more comprehensive characterization of the fragility or robustness of statistical significance specific to phase III oncology trials; to investigate multiple methods of evaluating SIFI; to compare the effects of different survival statistical models on SIFI; and to determine whether alternative approaches to clinical trial interpretation are needed.

## METHODS

We performed a meta-epidemiological analysis of phase III oncology trials identified from ClinicalTrials.gov in February 2020 with no date limitations. Only 2-arm superiority trials were included in this study. Primary endpoints (PEPs) that were not published, were not time-to-event, or lacked a Kaplan-Meir plot with a number-at-risk table were excluded. Screening with these criteria yielded 337 trials evaluable for the reconstruction of individual patients’ survival data. Methods of manual survival-data reconstruction using WebPlotDigitizer (Austin, TX) have been reported previously.^17,18^ The reconstruction quality was defined as the absolute value of the natural logarithm of HR_recon_/HR_reported_, where HR_recon_ was the point estimate of the hazard ratio (HR) for the reconstructed data determined by Cox regression, and HR_reported_ was the point estimate of the HR reported in the trial publication. Reconstructions in which this value was greater than 0.1 were excluded. Trials with proportional hazards violations in the PEP were also excluded, because the Cox proportional hazards model, used for the SIFI estimation, is unreliable in this setting.^19^ After application of these criteria, 230 trials were eligible for inclusion in the study and further analysis, as reported previously.^17^ Trial-level features were recorded in a standardized database. Enrollment was defined as the number of patients in the PEP analysis. Surrogate endpoints were defined as disease-related endpoints attempting to represent overall survival, consistent with other publications.^20^ United States’ Food and Drug Administration (FDA) approvals were evaluated as reported previously.^21^ Institutional review board approval was not needed because the data were publicly available. This report conforms to the modified Preferred Reporting Items for Systematic Reviews and Meta-Analyses (PRISMA) for meta-epidemiological studies.^22^

The fragility of each trial was estimated using R v.4.4.2 (Vienna, Austria) with previously published methods, and the code is included in the **Supplement**.^16^ SIFI values were estimated by counting the number of patients that needed to be iteratively flipped between treatment arms to change the original statistical significance interpretation of the PEP Cox proportional hazards regression (i.e., from positive to negative or vice versa). Statistical significance was defined uniquely for each trial as the threshold set by the trial. Flipping of the best (longest) survivors from the experimental arm to the control arm to calculate SIFI_B_ was used for the main analysis. Sensitivity analyses were performed by flipping the worst (shortest) survivors from the control arm to the experimental arm (SIFI_W_), and flipping of the median survivors from the control arm to the experimental arm (SIFI_M_). SIFI counts were computed using R v4.4.2 (Vienna, Austria). After the absolute value of the SIFI count was calculated, the index was normalized as a percentage of the total number of participants evaluated in the PEP analysis. Based on prior work, we defined severe fragility as being present when the SIFI was less than or equivale to 1% of the study enrollment.^16^

To explore the effects of the survival model on SIFI, SIFI, as conventionally estimated by Cox proportional hazards regression, was compared to SIFI estimated by three alternative survival models: restricted mean survival time (RMST), MaxCombo2, and MaxCombo3. RMST models estimate the difference in mean survival between treatment arms by integrating the area under the survival curve until the truncation time *τ*, defined here as the earlier of the last observed event from either arm.^19^ MaxCombo2 incorporates the Fleming–Harrington log-rank statistic plus a weighted log-rank statistic for late separation of curves, and MaxCombo3 provides an additional weighting to MaxCombo2 to account for diminishing treatment effects.^23^

Continuous variables were summarized by median and interquartile range (IQR). Correlations between the SIFI and the initial *P* value were estimated using the Spearman’s rank correlation coefficient. The chi-square test was used to test the association between categorical variables. The associations between trial-level features and the SIFI were evaluated using the Mann-Whitney U test. Binary logistic regressions tested the association between trial-level features and severe fragility to estimate odds ratios (OR). Two-sided *P* values with 95% CIs were calculated using SAS v9.4 (Cary, NC), and significance was defined as *P* < 0.05. Plots were created using Prism v10 (La Jolla, CA).

## RESULTS

The study included 230 trials, published from 2005 to 2020 and enrolling 184,752 patients in total (**Table S1**). Most trials evaluated surrogate PEPs (n=140, 61%). The PEP was positive (i.e., the experimental arm was interpreted as demonstrating superiority to the control arm) in 120 trials (52%), and led to FDA approving the experimental therapy tested in 82 trials (36%).

In the overall dataset, the median SIFI_B_, SIFI_W_, and SIFI_M_ counts were 8 patients (IQR, 4 to 19 patients), 11 patients (IQR, 6 to 20 patients), and 17 patients (IQR, 9 to 30 patients), respectively (**Figure 1A**). As percentages of the number of patients studied in the PEP analyses, the median SIFI_B_, SIFI_W,_ and SIFI_M_ percentage were 1.4% (IQR, 0.7% to 3%), 1.9% (IQR, 0.9% to 3.8%), and 3% (IQR, 1.3% to 6.0%), respectively (**Figure 1B**). The SIFI values estimated by each method were well-correlated, especially for the SIFI_W_ and SIFI_M_ values (**Figure 2**). Severe fragility (i.e., SIFI percentage *≤* 1% of the study enrollment) was detected in 87 trials (38%) per the SIFI_B_ values, 68 trials (30%) per the SIFI_W_ values, and 49 trials (21%) per the SIFI_M_ values.

**Figure 1.**
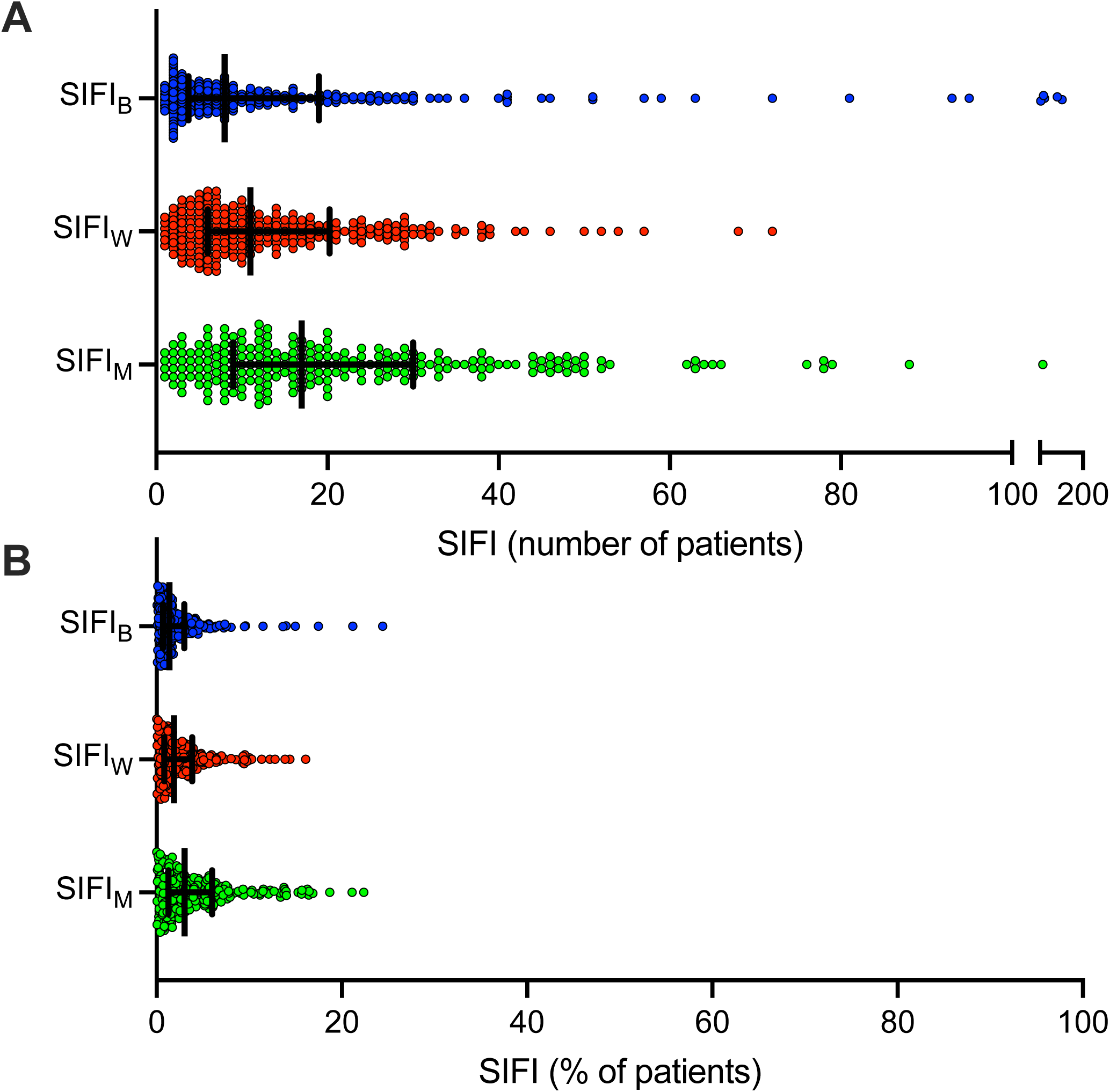
The survival-inferred fragility index (SIFI) for the primary endpoints of phase III oncology trials according to SIFI_B_, SIFI_W_, and SIFI_M_. (A) The absolute numbers of patients and (B) the percentages of the number of patients studied in the primary endpoint analyses are shown. Bars represent medians and interquartile ranges.

**Figure 2.**
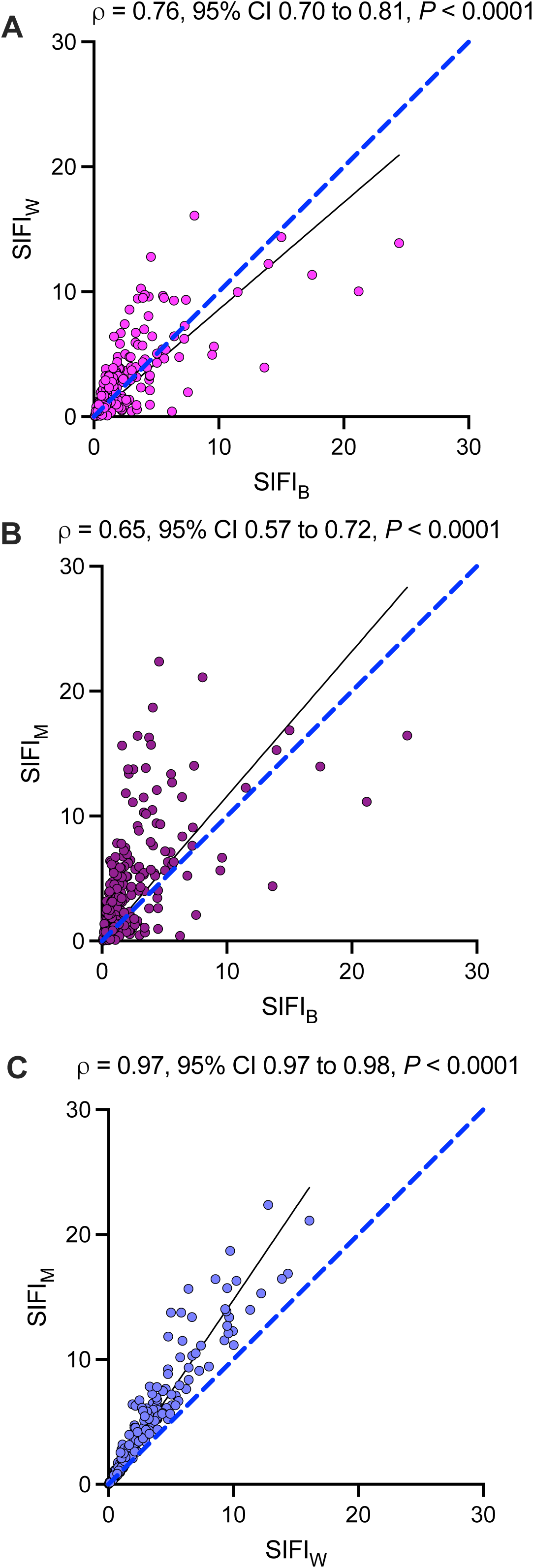
Different methods of calculating survival-inferred fragility index (SIFI) percentages provide highly concordant results. The Spearman’s rank correlation coefficient is shown. In each figure, the solid line is the best fit univariable regression. The dashed line is the line of identity.

Trials with overall survival PEPs were significantly more fragile (i.e., had lower SIFI percentages) than trials with surrogate PEPs (median SIFI_B_ percentages: 1.0% vs 1.8%, respectively; *P* < 0.0001) (**Figure 3A**). Similarly, compared with trials with surrogate PEPs, trials with overall survival PEPs were associated with greater odds of severe fragility, as defined by the SIFI_B_ value (OR, 2.33; 95% CI, 1.35 to 4.06; *P* = 0.003). This difference may have been related to the fact that trials claiming superiority appeared modestly less fragile (i.e., had higher SIFI values) than trials that did not claim superiority (median SIFI_B_ percentages: 1.9% vs 1.1%, respectively; *P* = 0.0002) because surrogate PEPs were more likely to result in superiority interpretations than overall survival PEPs (64% vs 30%, respectively; *P* < 0.0001 per the chi-square test) (**Figure 3B**). However, in the context of severe fragility, there was only a weak signal that initial statistical significance (i.e., claims of superiority) was associated with reduced odds of severe SIFI_B_ fragility (OR, 0.62; 95% CI, 0.36 to 1.06; *P* = 0.08). This association became inconclusive after adjusting for the potential confounding effects of PEP type on trial outcome (adjusted OR, 0.78; 95% CI, 0.44 to 1.37; *P* = 0.38).

**Figure 3.**
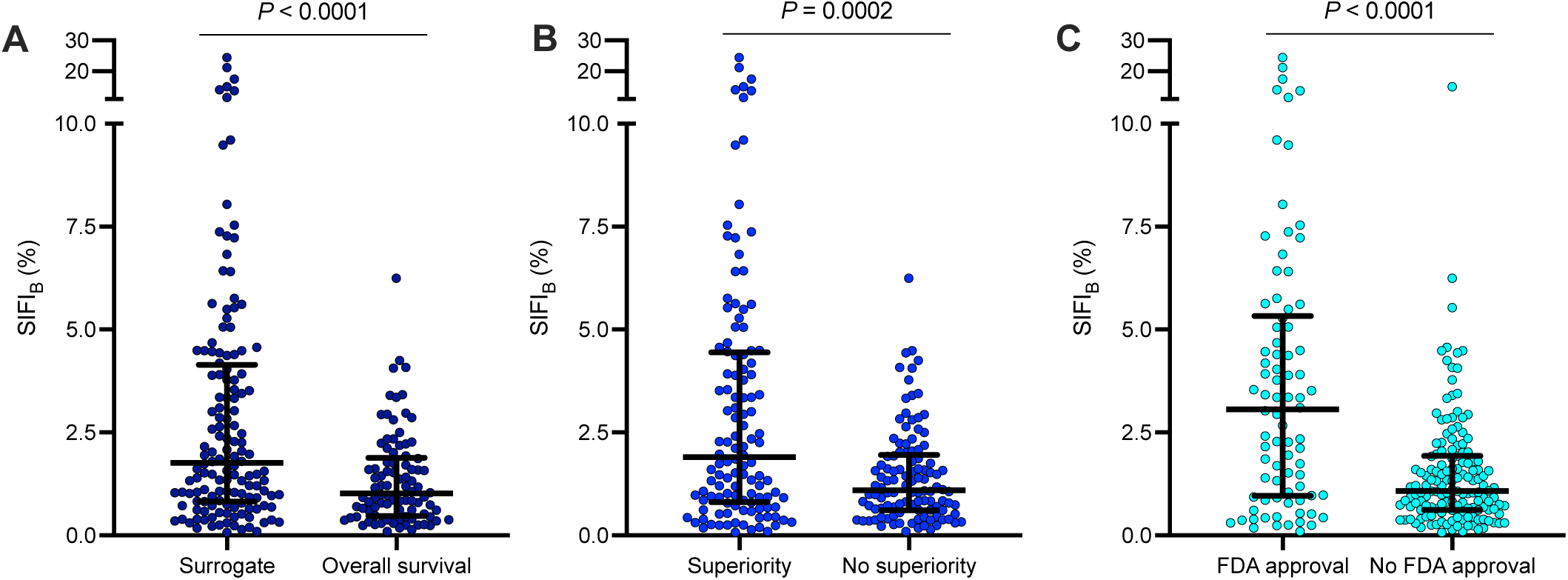
Trials claiming superiority, studying surrogate endpoints, and receiving FDA approval tend to have larger survival-inferred fragility indices (SIFI). The association of fragility, as measured by SIFI_B_, with trial-level features: (A) Primary endpoint type; (B) statistical significance; (C) Subsequent FDA approval. *P* by Mann-Whitney U test. Bars represent medians and interquartile ranges.

Interestingly, despite these findings for the interpretation of *P*, the value of *P* itself appeared closely associated with severe SIFI_B_ fragility (-log_10_(*P*): OR, 0.73; 95% CI, 0.63 to 0.83; *P* < 0.0001) and after adjustment for PEP type (adjusted OR, 0.75; 95% CI, 0.64 to 0.86; *P* = 0.0001). This result was consistent when fragility was estimated for each SIFI approach (**Figure S1**).

On the other hand, the differences in fragility were more pronounced in the comparison of trials that did versus did not lead to FDA approval. The median SIFI_B_ percentage of trials leading to FDA approval was 3.1%, and the median SIFI_B_ percentage of trials not leading to FDA approval was vs 1.1% (*P* < 0.0001) (**Figure 3C**). Trials with FDA approval were associated with lower odds of severe SIFI_B_ fragility than trials without subsequent FDA approval (OR, 0.47; 95% CI, 0.26 to 0.83; *P* = 0.01). This association persisted after adjustment for PEP type (adjusted OR, 0.53; 95% CI, 0.29 to 0.96; *P* = 0.04). There was also a significant relationship between FDA approval and severe fragility as estimated using SIFI_W_ values (adjusted OR, 0.52; 95% CI, 0.27 to 0.96; *P* = 0.04), although the effect was weaker for severe fragility as estimated using SIFI_M_ values (aOR, 0.59; 95% CI, 0.28 to 1.17; *P* = 0.14). Thus, although the SIFI percentages were low across the full dataset, including among trials claiming superiority, the FDA approval process did appear to select for findings that appeared, on the whole, notably less fragile (i.e., the median fragility of trails receiving approval was approximately one third that of trials not receiving approval). Other characteristics were not associated with fragility, such as whether or not the trial was an immunotherapy study (median SIFI_B_ percentage, 1.47% [IQR, 0.60% to 4.0%] for immunotherapy trials vs 1.40% [IQR, 0.7% to 2.9%] for trials testing other approaches; *P* = 0.80).

The effects of different survival models on estimating the SIFI values are shown in **Figure 4**. The SIFI_B_ values were highest (i.e., least fragile) when estimated by the RMST model (*P* < 0.0001 vs Cox), and lowest (i.e., most fragile) when estimated by the MaxCombo approaches (*P* < 0.0001 and *P* = 0.03 for MaxCombo2 and MaxCombo3, respectively, vs Cox) (**Figure 4A**). The inverse was demonstrated for the SIFI_W_ values, where MaxCombo2 and MaxCombo3 were found to be least fragile, when compared with Cox, and RMST most fragile (**Figure 4B**).

**Figure 4.**
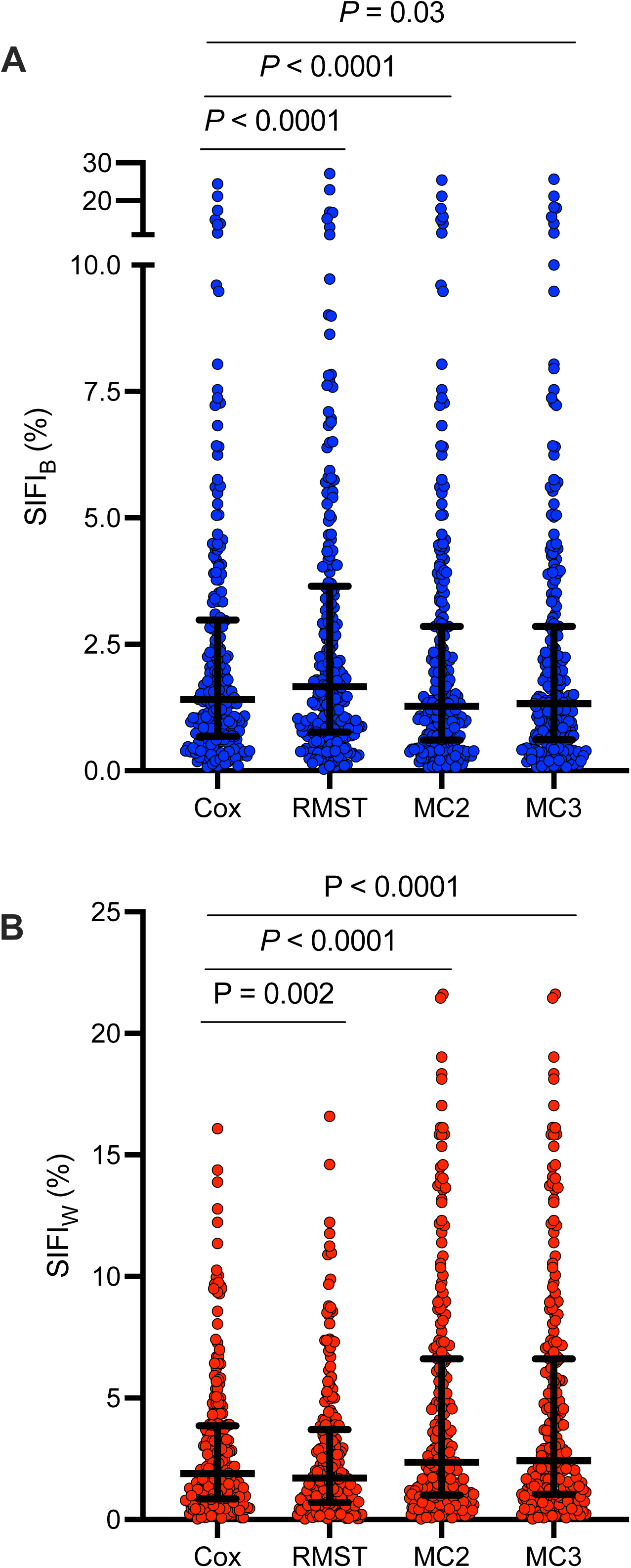
The survival-inferred fragility index (SIFI) estimates are influenced by the underlying survival model. (A) SIFI_B_ and (B) SIFI_W_ estimated using Cox regression, restricted mean survival time (RMST), MaxCombo2 (MC2), and MaxCombo3 (MC3).

## DISCUSSION

In this study, we estimated the fragility of phase III oncology trial results by leveraging reconstruction techniques to assemble a uniquely comprehensive dataset of 184,752 individual patient outcomes from 230 trials. We found evidence of fragility as estimated by the SIFI approach in most trials; the median SIFI count was 8 patients, or a median percentage of the study enrollment of 1.4%. Severe fragility was identified in 87 (38%) trials. Taken together, our results provide novel and quantitative insights into the problems of using statistical significance to interpret the results of phase III oncology trials. Alternative approaches to trial interpretation that do not rely solely on statistical significance are urgently needed.

We found that severe fragility, while related to the initial *P* value as a continuous parameter, does not appear to be closely related to whether the initial *P* value was deemed significant or not. In other words, both “positive” and “negative” trials, as defined by statistical significance, are susceptible to fragility, and a small series of changes to individual outcomes may readily reverse the trial interpretation in either direction. This finding is not necessarily surprising from a conceptual standpoint; the information provided by *P* values of 0.049 and 0.051 is essentially identical.^24^ However, this empirical observation is concerning because it implies that the extent of both type I errors and type II errors are currently underestimated in phase III oncology trials.^17^ However, when *P* is viewed as a continuous statistic rather than a binary outcome, which is a more appropriate approach, *P* serves well as a marker of fragility.^3^ While trials that obtained FDA approval did seem to exhibit less fragility on average, suggesting the importance of the downstream regulatory approval process, fragility still remained problematic even for this subset of trials, as the median SIFI value was 3%.

In our study, different approaches to estimating SIFI reveal the strengthens and weaknesses of the concept of fragility. We found strong concordance between SIFI estimated according to flipping the best survivor, the worst (shortest) survivor, and the median survivor. Moreover, fragility appears to be markedly influenced by the choice of the underlying survivor model. RMST analyses, which compare the area under the survival curve, appeared less fragile when the longest-term survivors were flipped between arms, consistent with expectations that late-occurring drops will hold less influence on area under the curve calculations. Conversely, MaxCombo models, which weight for late separation of the curves or late diminishing treatment effects, appeared more fragile when flipping the best survivors. We found the inverse to be the case when focusing SIFI on the earliest aspects of the survival curve in the SIFI_W_ models. Thus, the underlying data from an individual trial and the underlying survival model could considerably influence the results of fragility estimations, and each should be interpreted as offering distinct and unique information on the behavior of the underlying survival data.

Alternative approaches to clinical trial interpretation beyond statistical significance are urgently needed, and ultimately, the regulatory process, as perhaps implied by our data, appears most strongly poised to affect such change.^25^ The European Society of Medical Oncology and the American Society of Clinical Oncology have proposed that evaluations of treatment effect superiority should include assessments of effect size.^26–28^ Effect sizes describe the relative magnitude of benefit to patients and adds important clinical meaning to the interpretation of survival data.^29^ Notably, because *P* values do not convey effect sizes (although they are often perceived in this way), treatment effects with very low *P* values (e.g., *P* < 0.001) may be associated with only marginal effect sizes.^7,30^ Quantifying the probability of achieving clinically meaningful effect sizes in phase III oncology trials is a highly valuable strategy for trial interpretation, and would complement the use of statistical significance.^17,31^ Lastly, it is essential to not only consider the challenges of trial interpretation, the generalizability of the trial, and caveats of trial design, but to also take into account the characteristics and needs of individual patients—such as patient values and patient-specific risks—when making clinical decisions.^32,33^

Caution is warranted when interpreting the present study for several reasons. This study’s findings are subject to the limitations of reconstructed survival data.^18^ Although trial-specific significance levels were used to mimic the conditions of the original studies, all reconstructed regressions were univariable. In contrast, most phase III oncology trials use multivariable regressions for the PEP because multivariable regressions have greater power and efficiency than univariable regressions.^34,35^ Thus, the SIFI modeling approach may overestimate fragility for trials that were initially statistically significant, because lower *P* values, which reflect more information than higher *P* values, may be obtained when strongly prognostic covariates are included in the PEP regression model.^35^ To reduce this bias from this risk, our analysis excluded studies where the HR based on reconstructed data differed from the original HR by more than 0.1 on the logarithmic scale. Like any single summary measure, the fragility index has limitations, and we do not suggest that it can provide a stand-alone alternative to the p-value for making decisions regarding treatment efficacy.^16^ There is no consensus approach among researchers regarding the definitions of SIFI and severe fragility, and so we used multiple approaches to examine SIFI. Despite these limitations, we propose that the fragility index provides an intuitive metric for shedding light on the instability of clinical trial results.

Although studying SIFI is ultimately *in silico*, one of the key strengths of this study, compared to a pure-simulation study, is that actual patient outcomes were used to estimate fragility. Thus, our findings here have direct and immediate relevance to phase III oncology trials, which frequently change the standard of care. Building on a growing literature conveying the limitations of statistical significance criteria, this study provides new, quantitative, and easily understandable insights into the severe fragility of many late-phase trials. To improve the reliability of the evidence generated in oncology trials, alternative strategies for interpreting clinical trials beyond statistical significance are urgently needed.

## Data availability

Deidentified reconstructed data for individual patients are available at the online repository Figshare and accessible via: https://figshare.com/articles/dataset/Reconstructed_survival_data_from_Phase_3_oncol<u>ogy_trials/26103268</u>. Code for this study is included in the supplement.

## Data Availability

Deidentified reconstructed data for individual patients are available at the online repository Figshare and accessible via: https://figshare.com/articles/dataset/Reconstructed_survival_data_from_Phase_3_oncology_trials/26103268. Code for this study is included in the supplement.

## Acknowledgments

We thank Laura Russell, scientific editor in The University of Texas MD Anderson Cancer Center’s Research Medical Library, for editing the manuscript.

## Previous presentations

Portions of this study were previously presented in October 2024 at the American Society for Radiation Oncology Annual Meeting in Washington DC.

## Support

This work was supported in part by the National Institutes of Health/National Cancer Institute through MD Anderson’s Cancer Center Support Grant P30CA016672. Pavlos Msaouel and Ethan Ludmir are recipients of the Andrew Sabin Family Foundation Fellowship.

## Authors’ disclosures of potential conflicts of interest

Alexander Sherry reports honoraria from Sermo. Pavlos Msaouel reports honoraria for scientific advisory board membership for Mirati Therapeutics, Bristol-Myers Squibb, and Exelixis; consulting fees from Axiom Healthcare; non-branded educational programs supported by DAVA Oncology, Exelixis, and Pfizer; leadership or fiduciary roles as a Medical Steering Committee Member for the Kidney Cancer Association and as a Kidney Cancer Scientific Advisory Board Member for KCCure; and research funding from Regeneron Pharmaceuticals, Takeda, Bristol-Myers Squibb, Mirati Therapeutics, and Gateway for Cancer Research (all unrelated to this manuscript’s content). Zachary McCaw reports employment at Insitro (unrelated to this manuscript’s content). Tomer Meirson reports consulting fees from Purple Biotech. No other authors report any conflicts of interest.

## Conception and design

Alexander Sherry, Yufei Liu, Pavlos Msaouel, Timothy Lin, Jonathan Ofer, David Bomze, Zachary McCaw, Tomer Meirson, Ethan Ludmir.

## Collection and assembly of data

Alexander Sherry, Timothy Lin, Alex Koong, Christine Lin, Joseph Abi Jaoude, Roshal Patel, Ramez Kouzy, Molly El-Alam, Avital Miller, Ethan Ludmir. **Financial support:** Ethan Ludmir. **Software and code:** Alexander Sherry, Yufei Liu, Timothy Lin, Mohannad Owiwi, Jonathan Ofer, David Bomze, Tomer Meirson, Ethan Ludmir. **Data analysis and interpretation**: All authors. **Manuscript writing:** ADS wrote the first draft. All authors revised the paper for critical intellectual content. **Final approval of manuscript:** All authors. **Accountable for all aspects of work:** All authors

## Notes

### Funding Statement

This work was supported in part by the National Institutes of Health National Cancer Institute through MD Anderson Cancer Center Support Grant P30CA016672. Pavlos Msaouel and Ethan Ludmir are recipients of the Andrew Sabin Family Foundation Fellowship.

