## Supplement for "Survival-Inferred Fragility of Statistical Significance in Phase III Oncology Trials"

Sherry AD et al.

**Table S1**. Characteristics of the 230 included trials.

**Figure S1**. Fragility is lower in the setting of lower *P* values as a continuous statistic.

**Table S1**. Characteristics of the 230 phase III oncology trials included in this study.

| Characteristic | Value |
|---|---|
| Cancer stage and type, n (%) | |
| Solid nonmetastatic | 51 (22) |
| Solid metastatic | 141 (61) |
| Hematologic | 38 (17) |
| Cancer site, n (%) | |
| Breast | 45 (20) |
| Gastrointestinal system | 41 (18) |
| Genitourinary system | 35 (15) |
| Blood | 38 (17) |
| Thoracic system | 40 (17) |
| Other[a] | 31 (13) |
| Treatment modality, n (%) | |
| Systemic therapy | 224 (97) |
| Immunotherapy | 17 (7) |
| Local therapy | 6 (3) |
| Cooperative group sponsorship, n (%) | 53 (23) |
| Industry-funded trial, n (%) | 202 (88) |
| Publication year, median (IQR) | 2015 (2013 – 2017) |
| PEP, n (%) | |
| Overall survival | 90 (39) |
| Surrogate | 140 (61) |
| Number of patients in the PEP, median (IQR) | 594 (406 – 861) |
| PEP outcome, n (%) | |
| Positive | 120 (52) |
| Negative | 110 (48) |
| Trial results led to FDA approval of treatment, n (%) | 82 (36) |

[a] Other disease sites include: central nervous system, endocrine, gynecologic, head and neck, and skin.

Abbreviations: IQR (interquartile range); PEP (primary endpoint); FDA (US Food and Drug Administration).

**Figure S1**. Fragility is lower in the setting of lower *P* values as a continuous statistic. Spearman's rank correlation coefficient is shown. In each figure, the solid line is the best-fit univariate regression.

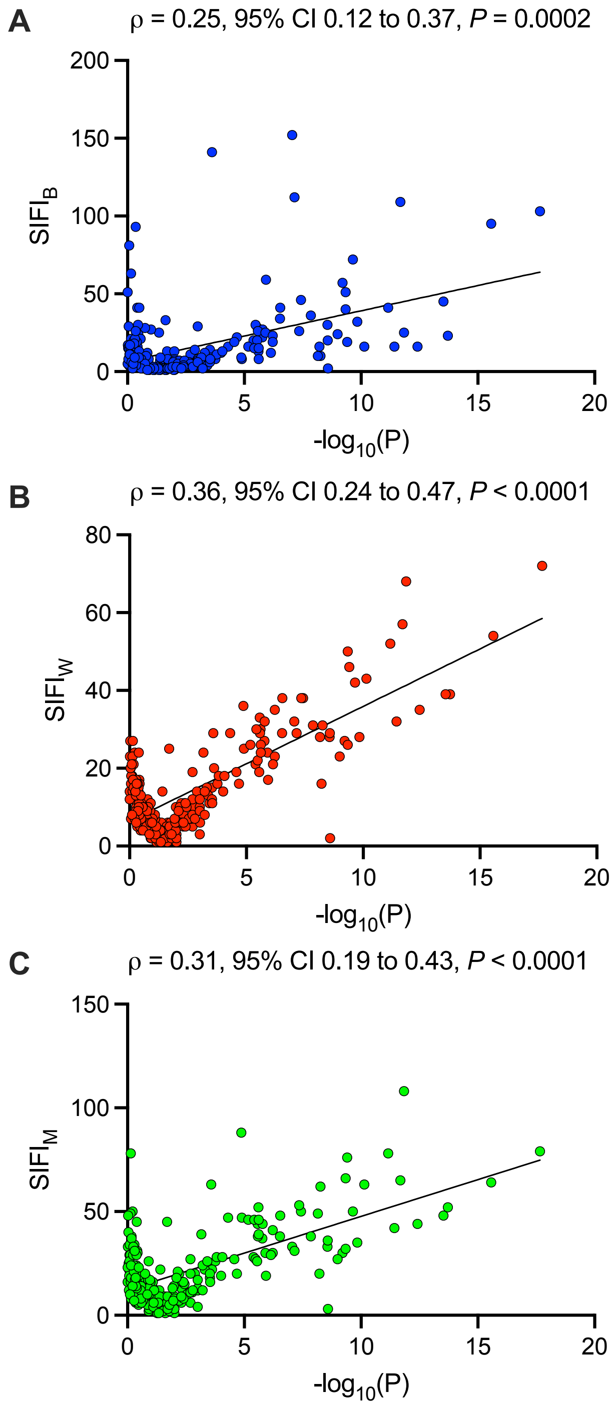
