## Supplemental code for "Survival-Inferred Fragility of Statistical Significance in Phase III Oncology Trials"

```
#@@@@@@@@@@@@@@@@@@@@@@@@@@@@@@@@@@@@@@@@@@@@@@@@@
# Basic SIFI function
#@@@@@@@@@@@@@@@@@@@@@@@@@@@@@@@@@@@@@@@@@@@@@@@@@
sifi <- function(sv_data, treatment_arm = NULL,  # 'sv_data' should contain three columns:
(1) time, (2) status, (3) arm
                 operation = c("flip","clone"),  # Flip or clone the best/worst responder
                 direction = c("best","worst","median"),  # Use the best (longest) the
worst (shortest) survivor or the median survivor from the control group
                 pval_thr = 0.05,                # this is a new addition - the threshold
to reject the null hypothesis
                 cols = c("#0754A0","#F12A29"),  # color of KM curves
                 stat_test = c("logrank","wald"),  # test used to calculate p-value in
each iteration
                 agnostic = F,   # Agnostic determination of experimental vs reference
group (based on the lower HR)
                 plot_iteration = F, file_iteration = NA){

  require(dplyr)
  require(survival)

  # Evaluate input
  operation <- match.arg(operation)
  direction <- match.arg(direction)
  stat_test <- match.arg(stat_test)

  # Prepare data
  if(all(colnames(sv_data)[1:3] == c('arm','time','status'))) sv_data =
sv_data[,c('time','status','arm')] # re-order columns
  names(sv_data) <- c("time","event","arm")
  sv_data$arm <- as.factor(sv_data$arm)
  sv_data$id <- 1:nrow(sv_data)
  count <- 0 ; flag <- T

  # If the treatment arm wasn't defined, we use the agnostic approach,
  # by assigning the group that shows benefit (HR < 1) as the experimental group
regardless of signifiance
  if((length(treatment_arm) == 0) | agnostic){
    sv_cox <- coxph(Surv(time, event, type = "right") ~ arm, data = sv_data)
    treatment_arm <- ifelse(sv_cox$coefficients < 0, yes = levels(sv_data$arm)[2], no =
levels(sv_data$arm)[1])
  }

  # Original count
  n_arms <- table(sv_data$arm)

  #@@@@@@@@@@ WE NOW HAVE 4 OPTIONS (2x2)
  # 1) Re-designate the best responder  (longest time)  from experimental to control group
  # 2) Re-designate the worst responder (shortest time) from control to experimental group
  # 3) Flip that responder, 4) Clone that responder
  #@@@@@@@@@@

  # Plot the iteration to PDF if we have filename, otherwise to Viewer
  if(plot_iteration & !is.na(file_iteration)){
    cairo_pdf(file_iteration, width = 11.69, height = 8.27, onefile = T)
    par(mfrow = c(3,4))
  }

  # Initialize while loop
  while(flag){

    # Option A: Calculate p-value of log-rank test (default)
    if(stat_test == "logrank"){
      sdf <- survdiff(Surv(time, event, type = "right") ~ arm, data = sv_data)
      pval <- 1 - pchisq(sdf$chisq, length(sdf$n) - 1)  # Log-rank p-value directly from
```

```
survdiff
    }

    # Option B: Calculate p-value of Wald's test
    if(stat_test == "wald"){
      cxp <- coxph(Surv(time, event, type = "right") ~ arm, data = sv_data)
      pval <- summary(cxp)$coefficients[5]
    }

    #@@@@@@@@@@@ NEGATIVE SIFI, if in the first iteration the p-val is insignificant
    # calculate the negative SIFI, i.e. try to get it from non-significant to significant
    if(count == 0 & pval > pval_thr){
      # Dump parameters
      count_neg <- neg_sifi(sv_data = sv_data[ , 1:3], treatment_arm = treatment_arm,
                            operation = operation, direction = direction,pval_thr =
pval_thr,
                            cols = cols, stat_test = stat_test,
                            agnostic = agnostic,
                            plot_iteration = plot_iteration, file_iteration =
file_iteration)
      return(count_neg)
    }
    #@@@@@@@@@@@

    if(plot_iteration){
      # Build survival model, run COXPH, and calculate HR
      sft <- survfit(Surv(time, event, type = "right") ~ arm, data = sv_data)
      cxp <- coxph(Surv(time, event, type = "right") ~ arm, data = sv_data)
      hr <- summary(cxp)$conf.int[c(1,3,4)]

      # Create labels
      hr_lab <- paste0("HR = ", sprintf("%1.2f", hr[1]), " (",
                       sprintf("%1.2f", hr[2]), ",", sprintf("%1.2f", hr[3]), ")")
      pval_lab <- ifelse(pval < 0.001, yes = "p < 0.001", no = paste0("p = ",
sprintf("%1.3f", pval)))

      plot(sft, col = cols, lwd = 1.5,
           xlab = "", ylab = "% Survival", cex.lab = 1.0,
           main = paste0("Iteration #", count,
                         "\n", hr_lab,
                         "\n", pval_lab),
           mark.time = T, mark = "|" , cex = 0.7,
           xaxt = "n", yaxt = "n", frame.plot = FALSE)

      # Plot axes
      axis(side = 1, pos = 0) ; abline( h =0)
      title(xlab = "Time", line = 2.5, cex.lab = 1.0)
      axis(side = 2, at = seq(0, 1, 0.2), labels = seq(0, 1, 0.2)*100, las = 2)

      bounds <- par("usr")

      # Plot legend
      legend("bottomleft", legend = c("Experimental","Control"), col = rev(cols), bty =
"n", lwd = 1.5)

      # Add strategy (operation-direction)
      par(xpd = TRUE)   # Prevents clipping

      text(bounds[2], bounds[4]-0.05, adj = c(1,1), cex = 1.0,
           label = paste0(stringi::stri_trans_totitle(paste0(operation, " ", direction, "
responder")),
                          ifelse(direction == "best", yes ="\nExperimental --> Control",
no = "\nControl --> Experimental")))

      # If there is already a best/worst responder that was flipped/cloned (from previous
```

```
iteration), then plot it
      if(exists("jd")){
        points(jd[1], jd[2], type = "b", pch = 19, col = "red3", cex = 1.5)
        text(jd[1], jd[2]-0.05, adj = c(0.5,1), cex = 1.0, col = "red3",
             label = paste0("Just\n", ifelse(operation == "flip", "Flipped", "Cloned"), "
(", responder$event, ")"))
      }

    }

    # If we reached NON-significance, then we're done and return SIFI, else re-
deisgnate/add clone
    if(pval > pval_thr){
      if(plot_iteration & !is.na(file_iteration)) {
        dev.off()
      } # Shut down device we have a filename

      if(direction == "best") {
        write.csv(sv_data, file_4)
      }
      if(direction == "worst") {
        write.csv(sv_data, file_5)
      }
      if(direction == "median") {
        write.csv(sv_data, file_6)
      }
      return(count)
    } else {
      count <- count + 1
    }

    # Three options:
    # (1) Re-designate BEST responder from the experimental to the control group (either
event or censored)
    if(direction == "best") responder <- sv_data %>% filter(arm == treatment_arm) %>%
arrange(time) %>% tail(1)

    # (2) Re-desginate WORST responder from the control group to the experimental group
(must be event)
    if(direction == "worst") responder <- sv_data %>% filter(arm != treatment_arm & event
== 1) %>% arrange(time) %>% head(1)

    # (3) Re-desginate Mean responder from the control group to the experimental group
(must be event)
    if(direction == "median") responder <- sv_data %>% filter(arm != treatment_arm & event
== 1) %>% median.survivor()


    if(plot_iteration){
      # Calculate time of new responder
      s <- summary(sft, times = responder$time, extend = TRUE)  # Otherwise we get: 'Error
in array(xx, dim = dd) : vector is too large'
      # 'sr' some responder, 'jd' just redesignated
      if(direction == "best"){
        sr <- c(responder$time, s$surv[paste0("arm=", treatment_arm) == s$strata])
        jd <- c(responder$time, s$surv[paste0("arm=", treatment_arm) != s$strata])
      }
      if(direction %in% c("worst",'median')){
        sr <- c(responder$time, s$surv[paste0("arm=", treatment_arm) != s$strata])
        jd <- c(responder$time, s$surv[paste0("arm=", treatment_arm) == s$strata])
      }

      # Add the point of the responder (event/censored in parentheses)
      points(sr[1], sr[2], type = "b", pch = 19, col = "green4", cex = 1.5)
      text(sr[1], sr[2]+0.05, adj = c(0.5,0), cex = 1.0, col = "green4",
```

```
          label = paste0(ifelse(direction == "best", "Best", "Worst"),"\nResponder", "
(", responder$event, ")"))

      par(xpd = FALSE)   # Turn off
    }

    # Two options:
    # (3) Flip the responder from its original group to the other arm
    if(operation == "flip") sv_data[responder$id , "arm"] <- setdiff(levels(responder$arm)
, responder$arm)

    # (4) Clone the responder and ADD it to the other arm
    if(operation == "clone"){
      responder$id <- paste0(responder$id,"_clone")  # Add a tag
      responder$arm <- setdiff(levels(responder$arm) , responder$arm)  # Change to the
other arm
      sv_data <- rbind(sv_data, responder)  # Concatenate it to the original cohort
    }

    #@@@@@@@@@@@ KEEP IN MIND THAT IF WE CLONE A *CENSORED* INDIVIDUAL THE HR MAY NOT
CHANGE AND WE WILL GET STUCK IN A LOOP
    #@@@@@@@@@@@ THIS IS A SAFETY MECHANISM FOR THESE EXTREME CASES OR WHERE WE CAN'T REACH
NON-SIGNIFICANCE...
    if(count > min(n_arms)){
      if(plot_iteration & !is.na(file_iteration)) dev.off()  # Shut down device if we have
a filename
      return(NA)
    }
  }

}

#@@@@@@@@@@@@@@@@@@@@@@@@@@@@@@@@@@@@@@@@@@@@@@@@@@
# Negative SIFI function, called from 'sifi()'
#@@@@@@@@@@@@@@@@@@@@@@@@@@@@@@@@@@@@@@@@@@@@@@@@@@
neg_sifi <- function(sv_data, treatment_arm = NULL,  # 'sv_data' should contain three
columns: (1) time, (2) event, (3) arm
                     operation = c("flip","clone"),  # Flip or clone the best/worst
responder
                     direction = c("best","worst","median"),  # Use the best (longest) the
worst (shortest) survivor or median survivor from the control group
                     pval_thr = 0.05,                # this is a new addition - the
threshold to reject the null hypothesis
                     cols = c("#0754A0","#F12A29"),  # color of KM curves
                     stat_test = c("logrank","wald"),  # test used to calculate p-value in
each iteration
                     agnostic = F,   # Agnostic determination of experimental vs reference
group (based on the lower HR)
                     plot_iteration = F, file_iteration = NA){

  require(dplyr)
  require(survival)

  # Evaluate input
  operation <- match.arg(operation)
  direction <- match.arg(direction)
  stat_test <- match.arg(stat_test)

  # Prepare data
  if(all(colnames(sv_data)[1:3] == c('arm','time','status'))) sv_data =
sv_data[,c('time','status','arm')] # re-order columns
  names(sv_data) <- c("time","event","arm")
  sv_data$arm <- as.factor(sv_data$arm)
  sv_data$id <- 1:nrow(sv_data)
  count <- 0 ; flag <- T
```

```
  # If the treatment arm wasn't defined, we use the agnostic approach,
  # by assigning the group that shows benefit (HR < 1) as the experimental group
regardless of signifiance
  if((length(treatment_arm) == 0) | agnostic){
    sv_cox <- coxph(Surv(time, event, type = "right") ~ arm, data = sv_data)
    treatment_arm <- ifelse(sv_cox$coefficients < 0, yes = levels(sv_data$arm)[2], no =
levels(sv_data$arm)[1])
  }

  # Original count
  n_arms <- table(sv_data$arm)

  #@@@@@@@@@@ WE NOW HAVE 6 OPTIONS (3x2)
  # 1) Re-designate the best responder  (longest time)  from experimental to control group
  # 2) Re-designate the worst responder (shortest time) from control to experimental group
  # 3) Re-designate the median responder from control to experimental group
  # a) Flip that responder, b) Clone that responder
  #@@@@@@@@@@

  # Plot to PDF if we have filename, otherwise to viewer
  if(plot_iteration & !is.na(file_iteration)){
    cairo_pdf(file_iteration, width = 11.69, height = 8.27, onefile = T)
    par(mfrow = c(3,4))
  }

  # Initialize while loop
  while(flag){

    # Option A: Calculate p-value of log-rank test (default)
    if(stat_test == "logrank"){
      sdf <- survdiff(Surv(time, event, type = "right") ~ arm, data = sv_data)
      pval <- 1 - pchisq(sdf$chisq, length(sdf$n) - 1)  # Log-rank p-value directly from
survdiff
    }

    # Option B: Calculate p-value of Wald's test
    if(stat_test == "wald"){
      cxp <- coxph(Surv(time, event, type = "right") ~ arm, data = sv_data)
      pval <- summary(cxp)$coefficients[5]
    }

    if(plot_iteration){
      # Build survival model, run COXPH, and calculate HR
      sft <- survfit(Surv(time, event, type = "right") ~ arm, data = sv_data)
      cxp <- coxph(Surv(time, event, type = "right") ~ arm, data = sv_data)
      hr <- summary(cxp)$conf.int[c(1,3,4)]

      # Create labels
      hr_lab <- paste0("HR = ", sprintf("%1.2f", hr[1]), " (",
                       sprintf("%1.2f", hr[2]), ",", sprintf("%1.2f", hr[3]), ")")
      pval_lab <- ifelse(pval < 0.001, yes = "p < 0.001", no = paste0("p = ",
sprintf("%1.3f", pval)))

      plot(sft, col = cols, lwd = 1.5,
           xlab = "", ylab = "% Survival", cex.lab = 1.0,
           main = paste0("Iteration #", count,
                         "\n", hr_lab,
                         "\n", pval_lab),
           mark.time = T, mark = "|" , cex = 0.7,   # Might be useful to keep for
illustration
           xaxt = "n", yaxt = "n", frame.plot = FALSE)

      # Plot axes
      axis(side = 1, pos = 0) ; abline( h =0)
```

```
      title(xlab = "Time", line = 2.5, cex.lab = 1.0)
      axis(side = 2, at = seq(0, 1, 0.2), labels = seq(0, 1, 0.2)*100, las = 2)

      bounds <- par("usr")

      # Plot legend
      legend("bottomleft", legend = c("Experimental","Control"), col = rev(cols), bty =
"n", lwd = 1.5)

      # Add strategy (operation-direction)
      par(xpd = TRUE)   # Prevents clipping

      text(bounds[2], bounds[4]-0.05, adj = c(1,1), cex = 1.0,
           label = paste0(stringi::stri_trans_totitle(paste0(operation, " ", direction, "
responder")),
                          ifelse(direction == "best", yes ="\nExperimental --> Control",
no = "\nControl --> Experimental")))

      # If there is already a best/worst responder that was flipped/cloned (from previous
iteration), then plot it
      if(exists("jd")){
        points(jd[1], jd[2], type = "b", pch = 19, col = "red3", cex = 1.5)
        text(jd[1], jd[2]-0.05, adj = c(0.5,1), cex = 1.0, col = "red3",
             label = paste0("Just\n", ifelse(operation == "flip", "Flipped", "Cloned"), "
(", responder$event, ")"))
      }

    }

    # If we reached YES-significance, then we're done and return SIFI, else re-
designate/add clone (mirror of positive SIFI)
    if(pval < pval_thr){
      if(plot_iteration & !is.na(file_iteration)) {
        dev.off()
      } # Shut down device we have a filename
      if(direction == "best") {
        write.csv(sv_data, file_4)
      }
      if(direction == "worst") {
        write.csv(sv_data, file_5)
      }
      if(direction == "median") {
        write.csv(sv_data, file_6)
      }
      return(count)
    } else {
      count <- count - 1   # Negative
    }

    # WE NOW MIRROR THE SAME APPROACH as positive SIFI
    control_arm <- setdiff(levels(sv_data$arm) , treatment_arm)

    # Two options:
    # (1) Re-designate BEST responder from the CONTROL to the EXPERIMENT group (i.e. the
mirror of positive SIFI) (either event or censored)
    if(direction == "best") responder <- sv_data %>% filter(arm == control_arm) %>%
arrange(time) %>% tail(1)

    # (2) Re-desginate WORST responder from the EXPERIMENTAL group to the CONTROL group
(i.e. the mirror of positive SIFI) (must be event)
    if(direction == "worst") responder <- sv_data %>% filter(arm != control_arm & event ==
1) %>% arrange(time) %>% head(1)

    # (3) Re-desginate Mean responder from the control group to the CONTROL group (i.e.
the mirror of positive SIFI) (must be event)
```

```
    if(direction == "median") responder <- sv_data %>% filter(arm != control_arm & event
== 1) %>% median.survivor()


    if(plot_iteration){
      # Calculate time of new responder
      s <- summary(sft, times = responder$time, extend = TRUE)  # Otherwise we get: 'Error
in array(xx, dim = dd) : vector is too large'
      # 'sr' some responder, 'jd' just redesignated
      if(direction == "best"){
        sr <- c(responder$time, s$surv[paste0("arm=", control_arm) == s$strata])  # mirror
of positive SIFI
        jd <- c(responder$time, s$surv[paste0("arm=", control_arm) != s$strata])  #
same...
      }
      if(direction %in% c("worst","median")){
        sr <- c(responder$time, s$surv[paste0("arm=", control_arm) != s$strata])  #
same...
        jd <- c(responder$time, s$surv[paste0("arm=", control_arm) == s$strata])  #
same...
      }

      # Add the point of the responder (event/censored in parentheses)
      points(sr[1], sr[2], type = "b", pch = 19, col = "green4", cex = 1.5)
      text(sr[1], sr[2]+0.05, adj = c(0.5,0), cex = 1.0, col = "green4",
           label = paste0(ifelse(direction == "best", "Best", "Worst"),"\nResponder", "
(", responder$event, ")"))

      par(xpd = FALSE)   # Turn off
    }

    # Two options:
    # (3) Flip the responder from its original group to the other arm
    if(operation == "flip") sv_data[responder$id , "arm"] <- setdiff(levels(responder$arm)
, responder$arm)

    # (4) Clone the responder and ADD it to the other arm
    if(operation == "clone"){
      responder$id <- paste0(responder$id,"_clone")  # Add a tag
      responder$arm <- setdiff(levels(responder$arm) , responder$arm)  # Change to the
other arm
      sv_data <- rbind(sv_data, responder)  # Concatenate it to the original cohort
    }

    #@@@@@@@@@@ KEEP IN MIND THAT IF WE CLONE A *CENSORED* INDIVIDUAL THE HR MAY NOT
CHANGE AND WE WILL GET STUCK IN A LOOP
    #@@@@@@@@@@ THIS IS A SAFETY MECHANISM FOR THESE EXTREME CASES OR WHERE WE CAN'T REACH
NON-SIGNIFICANCE...
    if(abs(count) > min(n_arms)){   # Absolute because 'count' is negative
      if(plot_iteration & !is.na(file_iteration)) dev.off()  # Shut down device we have a
filename
      return(NA)
    }
  }

}

#@@@@@@@@@@@@@@@@@@@@@@@@@@@@@@@@@@@@@@@@@@@@@@@@@@
# Calculate all SIFI strategies in one run
#@@@@@@@@@@@@@@@@@@@@@@@@@@@@@@@@@@@@@@@@@@@@@@@@@@
sifi_all <- function(sv_data, treatment_arm = NULL,  # 'sv_data' should contain three
columns: (1) time, (2) event, (3) arm
                     cols = c("#0754A0","#F12A29"),  # color of KM curves
                     stat_test = c("logrank","wald"),  # test used to calculate p-value in
each iteration
```

```
                     agnostic = F,   # Agnostic determination of experimental vs reference
group (based on the lower HR)
                     plot_iteration = F, file_prefix = NA){

  require(dplyr)

  # Evaluate input
  stat_test <- match.arg(stat_test)

  # Just need a data input, and run it 4 times
  #@@@@@@@@@@@ WE NOW HAVE 4 OPTIONS (2x2)
  # 1) Re-designate the best responder  (longest time)  from experimental to control group
  # 2) Re-designate the worst responder (shortest time) from control to experimental group
  # 3) Flip that responder, 4) Clone that responder
  #@@@@@@@@@@@

  # Create list to store SIFI
  sifi_all4 <- vector("list", 4)
  names(sifi_all4) <- expand.grid(operation = c("flip","clone"), direction =
c("best","worst")) %>%
    arrange(operation) %>%
    tidyr::unite("strategy", 1:2, remove = T) %>% pull(strategy)

  # Strategy ID
  strtgy <- 1

  # We run SIFI using the four different strategies
  for(op in c("flip","clone")){
    for(dr in c("best","worst")){
      sifi_all4[paste0(op, "_", dr)] <- sifi(sv_data = sv_data, treatment_arm =
treatment_arm,
                                             direction = dr, operation = op,
                                             cols = cols, stat_test = stat_test,
                                             agnostic = agnostic,   # Agnostic
determination of experimental vs reference group (based on the lower HR)
                                             plot_iteration = plot_iteration,
                                             file_iteration = ifelse(plot_iteration, yes =
paste0(file_prefix, "_strategy", strtgy, "_", paste0(op, "_", dr), ".pdf"), no = NA))
      strtgy <- strtgy + 1
    }
  }

  return(sifi_all4)
}

### Function to calculate the median survivor - or the first match near the median
median.survivor <- function(data) {
  index.med = which.min(abs(data$time - median(data$time)))
  data %>% slice(index.med)
}
```
